# Endovascular Treatment of Symptomatic Vasospasm and Delayed Cerebral Ischemia after Aneurysmal Subarachnoid Hemorrhage – a Systematic Review and Meta-analysis

**DOI:** 10.1101/2025.10.23.25338701

**Authors:** Michael Veldeman, Tobias Rossmann, Roel Haeren, Hanna Schenk, Rahul Raj, Charlotte Sabine Weyland

## Abstract

**Introduction:** Endovascular rescue treatment, including intra-arterial spasmolysis and/or transluminal balloon angioplasty, is widely used for symptomatic vasospasm and delayed cerebral ischemia (DCI) after aneurysmal subarachnoid hemorrhage (aSAH), yet its impact on functional recovery remains uncertain. We systematically reviewed and synthesized the evidence on patient-centered outcomes, exploring effects by follow-up time, intervention type, and clinical severity.

**Materials and methods:** Following a prospectively registered protocol (Research Registry: reviewregistry1466) and PRISMA guidance, we searched PubMed, EMBASE, and Web of Science (January 2000–December 2024; final update July 2025). Prospective and retrospective studies of adult aSAH patients receiving endovascular treatment for symptomatic angiographic vasospasm were included. Data extraction followed a standardized PICO framework, and study quality was assessed using the Newcastle–Ottawa Scale. Because reporting of angiographic resolution and DCI-related infarction was sparse or inconsistent, quantitative synthesis focused on dichotomized favorable functional outcome (e.g., mRS 0–2 or GOS good recovery). Single-arm meta-analyses of proportions were performed on the logit scale using random-effects generalized linear mixed models, with subgrouping by follow-up, intervention, and clinical severity.

**Results:** Thirty-nine studies (1,627 patients; 27 retrospective cohorts, 5 prospective cohorts, 5 case series, 2 randomized trials) met inclusion criteria; 38 contributed to meta-analysis. The pooled proportion of favorable functional outcomes was 0.55 (95% CI, 0.50–0.61) with substantial heterogeneity (I² ≈ 71%). Prespecified subgroup analyses by follow-up duration, intervention type, and baseline severity did not reveal significant differences. The two randomized trials reported conflicting short-term results with limited follow-up. Safety reporting was variable but generally acceptable for pharmacologic spasmolysis, while higher complication rates were occasionally observed with mechanical interventions.

**Conclusion:** Among patients with symptomatic vasospasm or DCI requiring endovascular rescue, approximately half achieve a favorable functional outcome. However, marked heterogeneity and reliance on predominantly observational data preclude firm conclusions regarding comparative effectiveness. Standardized multicenter randomized trials with harmonized definitions of eligibility, timing, outcomes, and adverse events are needed to clarify the therapeutic role and optimize patient selection for endovascular rescue after aSAH.

## Introduction

### Rationale

Delayed cerebral ischemia (DCI) remains the predominant complication contributing to poor neurological outcome following aneurysmal subarachnoid hemorrhage (aSAH).^1^ If left undiagnosed or when unsuccessfully treated, it may result in DCI-related infarction. Aside from the prophylactic use of oral nimodipine, no therapeutic intervention to date has consistently demonstrated a reduction in DCI incidence or severity nor contributed to an improvement in long-term clinical outcomes.^2,3^ Cumulative observational data suggest that induced hypertension, achieved through vasopressor infusion, serves as a first-line intervention upon diagnosis of DCI. However, a randomized controlled trial (RCT) evaluating this approach failed to demonstrate clinical benefit and was terminated early due to hypertension-related complications.^4^ This study has faced criticism for employing excessively high blood pressure targets, which likely contributed to the elevated complication rate. Moreover, the trial was underpowered to evaluate functional outcomes meaningfully.

While our understanding of DCI pathophysiology has evolved beyond a sole focus on angiographic large-vessel vasospasm, this phenomenon is still believed to represent the final common pathway leading to ischemic decompensation in vulnerable brain tissue. Contributing mechanisms include impaired cerebral autoregulation, disrupted neurovascular coupling, microthrombosis, microvasospasm, as well as cortical spreading depolarizations and ischemia.^5^

Notably, pharmacologic interventions that successfully reduced angiographic vasospasm have consistently failed to improve functional outcomes, highlighting the complexity of DCI pathogenesis.^6^ In recent years, advances in endovascular microcatheter technology and the availability of intra-arterial antithrombotic, vasodilatory agents, and intra-vascular devices^7^ have enabled the emergence of endovascular treatment strategies for DCI. Intra-arterial spasmolysis, via the infusion of vasodilators, has shown promise in resolving diffuse and distal vasospasm^8^, while transluminal balloon angioplasty may induce more sustained vessel dilation, albeit limited to focal, proximal vasospasm^9^.

Both techniques have been reported across numerous observational studies. To date, only two small randomized controlled trials have compared endovascular therapy with conservative management, but differences in study design and outcome evaluation have resulted in inconclusive and conflicting findings.^10,11^

### Objectives

This systematic review and meta-analysis aim to synthesize the current evidence regarding the efficacy of endovascular treatment for DCI following aSAH, specifically in (1) improving functional clinical outcome (2) and reducing DCI-related infraction.

## Methods

### Protocol and Registration

The review protocol was developed in September 2022 and included the definition of inclusion and exclusion criteria, specification of the time frame for eligible publications, and the formulation of a comprehensive trial evaluation checklist. The review was prospectively registered in October 2022 in the Research Registry (registration ID: reviewregistry1466). This manuscript is written in accordance with the PRISMA (Preferred Reporting Items for Systematic Reviews and Meta-Analyses) statement for reporting systematic reviews and adheres to methodological recommendations provided by the Cochrane Collaboration.

### Eligibility Criteria

All prospective and retrospective studies evaluating the effect of intra-arterial treatment for DCI were considered for inclusion. The search period was limited to studies published between January 2000 and July 2025. Eligible studies had to include adult patients (age ≥18 years) who received endovascular treatment for symptomatic angiographic vasospasm, either through the intra-arterial administration of vasodilatory medication or via mechanical intervention using balloon or stent angioplasty (e.g. Comaneci device).

Studies were eligible if they were prospective (including randomized and non-randomized designs), retrospective, or case series comprising at least five patients. Pilot studies and preliminary reports involving ten or more patients were considered for inclusion. Feasibility or safety studies, technical notes, and study protocols were excluded. Studies were required to report on at least one of the following: angiographic resolution of vasospasm, radiologic evidence of DCI based on perfusion CT or transcranial Doppler, the occurrence of DCI-related infarction, or clinical outcomes. Given that preliminary database searches revealed only a limited number of studies incorporating a control group, the presence of a control group was not considered an absolute requirement for inclusion. However, where available, studies with a control arm were earmarked for pooled analysis, provided that outcomes were sufficiently homogeneous.

### Information Sources and Search Strategy

A comprehensive literature search was conducted using the PubMed, EMBASE, and ISI Web of Science databases. Searches were performed at three time points: an initial exploratory search in January 2023, and a final search upon completion of data extraction in July 2025. The search strategy utilized the following terms in various combinations: “cerebral vasospasm,” “vasospasm,” “delayed cerebral ischemia,” “delayed ischemic neurological deficit,” “subarachnoid hemorrhage,” “spasmolysis,” and “angioplasty.” Variants in British and American spelling were included (e.g., isch(a)emia and h(a)emorrhage) to ensure comprehensive retrieval.

In addition to database searches, the reference lists of all included articles were manually screened to identify further eligible studies. Searches were restricted to articles published in English, German, French, or Dutch. Studies in other languages were considered for translation if they included an English abstract meeting the inclusion criteria and if translation of the full text was feasible.

### Data collection

Data extraction was conducted using a standardized template based on the PICO framework (Population, Intervention, Control, and Outcomes). The following variables were recorded: patient demographics (including age and sex), radiological and clinical severity of hemorrhage, overall sample size, specific indications for endovascular treatment, prior medical therapy, the operational definition of DCI applied, endovascular treatment-related complications, systemic complications, clinical outcomes, radiological outcomes, and study design.

All titles and abstracts retrieved from the search were imported into the Rayyan systematic review platform (www.rayyan.ai). Eligibility assessment was independently performed by two investigators (MV, CSW). Disagreements were resolved through discussion until a consensus was achieved. Duplicate records were identified and removed prior to full-text screening.

### Risk of Bias Assessment

The risk of bias for non-randomized studies was assessed using the Newcastle–Ottawa Scale (NOS), a validated tool for evaluating the quality of observational studies included in meta-analyses ^12^. The NOS assigns a maximum of eight points across three domains: selection, comparability, and outcome assessment, with higher scores reflecting higher methodological quality. Each included study was independently scored by two investigators. Discrepancies between reviewers were resolved through discussion until consensus was reached. NOS scores were subsequently translated into qualitative categories (good, fair, or poor) based on the standards of the Agency for Healthcare Research and Quality (AHRQ) for grading the risk of bias in individual studies.

### Data Analysis and Summary Measures

We pooled single-arm proportions of favorable functional outcome using R (v4.4.0; RStudio 2024.12) and the *meta* package. Outcome scales (e.g., modified Rankin Scale, mRS; Glasgow Outcome Scale, GOS) and assessment time points were reviewed a priori to judge comparability. Conventional dichotomization points for favorable outcome (mRS (0-2), GOS (4-5)) were adopted. Pooling and subgrouping followed this appraisal. Angiographic vasospasm resolution and DCI-related infarction were planned outcomes but were not pooled because reporting was infrequent and inconsistent; the primary synthesis therefore focused on functional outcomes.

Analyses used the logit scale with random-effects models. Generalized linear mixed models (GLMM) were applied for the primary analysis and for subgroups by follow-up duration and intervention type, given their robustness for proportions including 0% or 100%. For the clinical-severity subgroup, we also fit inverse-variance models (method = “Inverse”) on the logit scale to enable display of study weights. Inverse-variance weights, larger for more precise studies, are shown in forest plots and sum to 100% per analysis.

Between-study heterogeneity was summarized with τ² and I² and tested with Cochran’s Q. Pre-specified subgroups were follow-up ≤90 vs >90 days, intervention type (intra-arterial spasmolysis, balloon angioplasty, combined), and clinical severity (≥40% vs <40% poor grade at presentation). Poor grade was commonly defined as either Hunt & Hess or WFNS grade of 4 and 5. Subgroup differences were assessed with chi-square tests under common-effect and random-effects assumptions. Results are presented as forest plots with study estimates, 95% CIs, and, when applicable, weights. Interpretation emphasizes random-effects estimates (α = 0.05).

Publication bias and small-study effects were assessed using contour-enhanced funnel plots with conventional significance contours at 10%, 5%, and 1% and Egger’s regression test applied to logit-transformed proportions. Visual asymmetry and test results were interpreted cautiously given the high between-study heterogeneity and the single-arm proportion setting. Funnel plots and Egger’s tests were only performed for syntheses with at least ten studies to avoid low power and inflated Type I error. A two-sided p-value of < 0.05 was considered indicative of statistical significance.

## Results

### Study selection

Our search strategy identified 3,178 studies across three databases (PubMed, EMBASE, and Web of Science), of which 197 articles remained before duplicate removal (n=150). Following title and abstract screening, 47 full-text articles were assessed for eligibility. Eight manuscripts were excluded as they did not fulfill the defined inclusion criteria after full-text analysis. Finally, 39 studies met the eligibility criteria and were included in the systematic review, with 38 studies contributing data to the meta-analysis. One study was excluded to avoid double-counting, as it entailed a secondary analysis of the same patient cohorts. The literature search procedure is depicted in **Fig. 1**.

**Figure 1.**
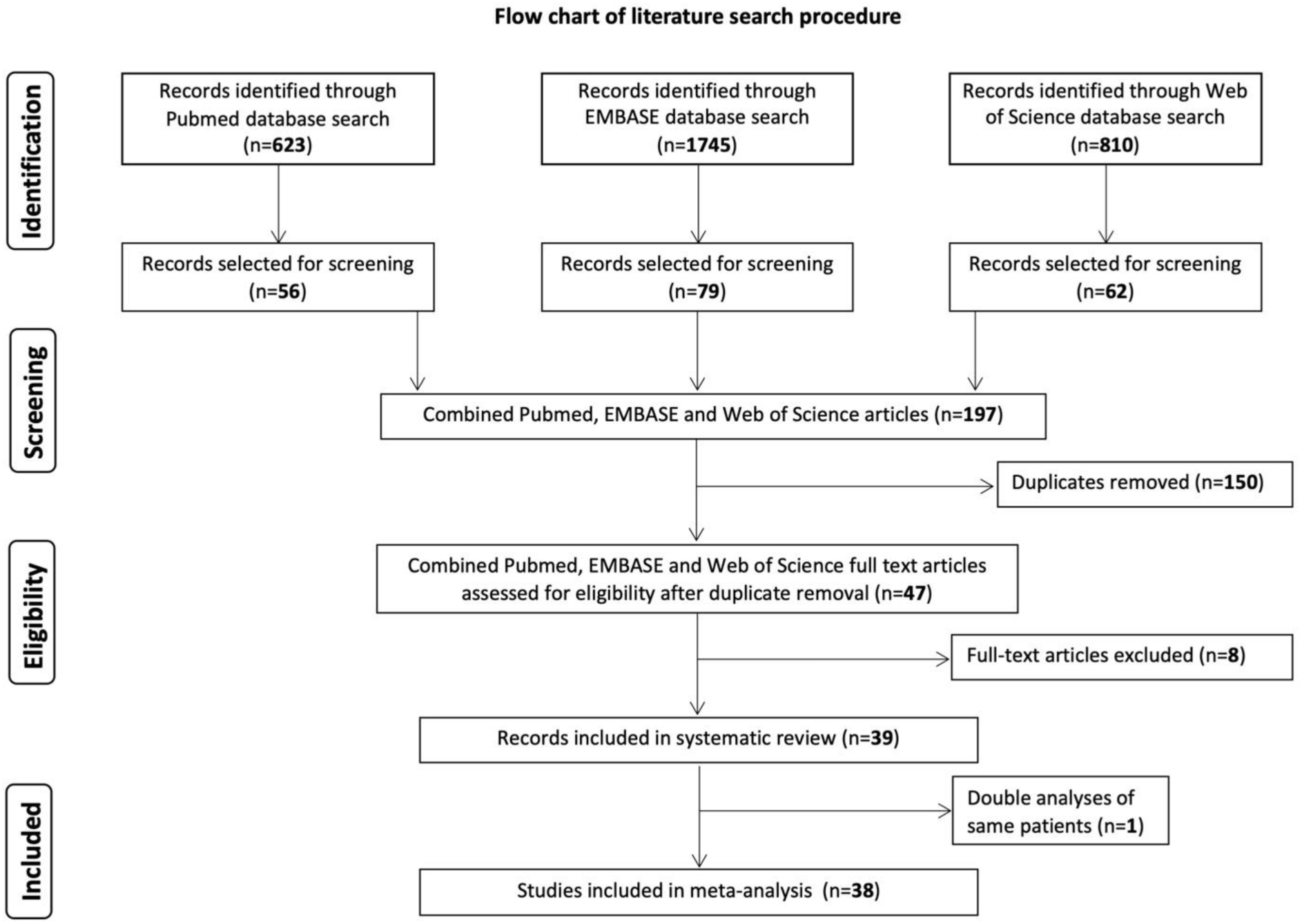
PRISMA flow diagram of literature search and study selection process. Systematic literature search across PubMed (n=623), EMBASE (n=1745), and Web of Science (n=810) databases. After duplicate removal and successive screening, 39 studies were included in the systematic review. Two studies with overlapping patient cohorts from the same center were identified ^9,47^; only one was included in the meta-analysis to avoid double-counting patients, resulting in 38 studies for quantitative analysis.

### Study characteristics

Of the 39 studies included in this systematic review, 27 were retrospective cohort studies (r-co) ^8,13–38^, five were case series (cs)^39–43^, five were prospective cohort studies (p-co)^9,44–47^, and two were randomized controlled trials (RCTs)^10,11^. Across all included studies, a total of 1,627 patients received endovascular treatment for delayed cerebral ischemia. The number of patients per study ranged from 5 to 202. A control group of some sort, was present in 13 studies. ^8,10,11,14,16,18,20,23,26,30,37,40,43^

### Risk of bias within and across studies

The methodological quality of the included studies, as assessed by the Newcastle–Ottawa Scale (NOS), varied considerably. Of the 39 studies, 11 were rated as having poor methodological quality, 20 as moderate, and only 8 studies were classified as high quality, according to the Agency for Healthcare Research and Quality (AHRQ) conversion standards. The average NOS score across all studies was 5.15 out of a possible 9, indicating a generally moderate to high risk of bias across the dataset.

The most frequent sources of bias were related to comparability between groups, which was inadequately addressed in 33 studies, and adequacy of follow-up, which was either unclear or insufficient in 23 studies. Selection bias was also common: 20 studies failed to report or implement appropriate selection of the non-exposed cohort, and 16 studies demonstrated limited representativeness of the exposed population. In contrast, outcome assessment and ascertainment of exposure were generally well reported, though still lacking in 6 and 5 studies, respectively. Taken together, the overall methodological limitations, especially in non-randomized studies, highlight the need for caution in interpreting pooled results. Detailed results of our interpretation and implementation of the RoB via the NOS tool are provided as **Supplemental Table 1**.

## Results of individual studies

### Randomized trials

To date, only two small RCTs have directly compared endovascular therapy to conservative medical management. A German trial randomized 34 patients with DCI (defined by perfusion deficit in perfusion MRI scanning), allocating 16 to receive endovascular treatment and 18 to standard care^10^. The trial was terminated prematurely due to a higher incidence of unfavorable outcomes, defined by the modified Rankin Scale (mRS) at six months, in the intervention arm. This cohort had not received prior induced hypertension, raising questions about treatment sequencing and patient selection.

In a second trial by Yindeedej et al., 68 patients were enrolled, with 36 undergoing intra-arterial nimodipine (IAN) treatment ^11^. This group demonstrated early neurological improvement, particularly in consciousness and motor function as measured by the Glasgow Coma Scale. However, the trial only included patients with an evaluable neurological examination, the clinical criteria for IAN administration were imprecise, and no follow-up beyond hospital discharge was provided.

### Prospective cohort studies

Small prospective cohorts showed short-term angiographic or physiologic responses with variable clinical impact. In 18 aSAH patients, balloon angioplasty or IAN produced transient perfusion and angiographic improvement.^44^ In 10 patients with abnormal invasive neuromonitoring, intra-arterial papaverine improved cerebral metabolism but the effect was short-lived.^45^ Among 25 patients with MRI-confirmed DCI, nimodipine spasmolysis or angioplasty was associated with a lower rate of DCI-related infarction.^46^ In a study by Weiss et al., about half of endovascularly treated patients reached functional independence at one year with low procedural complication rates. Bedside continuation of spasmolysis often increased vasopressor requirements, however, in this study, continuous spasmolysis in selected patients showed low complication rates and physiologic improvement, and 33% achieved favorable outcome at 3 months.^47^

### Retrospective cohort studies

Across retrospective cohorts, endovascular rescue commonly produced angiographic improvement, while clinical benefit varied. For example, Albrecht et al. reported 42.8% favorable outcome at 6 months and a low procedural complication rate of 4% across 241 interventions, with no ischemic or thromboembolic events recorded^14^. Early and frequent endovascular therapy was associated with lower infarction rates and better functional outcomes in a large pooled cohort analysis, although residual confounding is possible (Jabbarli 2019).^8^

Intra-arterial nimodipine (IAN) was the most frequently used regimen, occasionally continued bedside (CIAN) at the intensive care unit (ICU). Several cohorts described effective vasodilation and, in places, fewer infarcts or better outcomes, while others noted limited translation of angiographic gains into clinical benefit (Anthofer^16^, Dehdashti^17^, Goel^20^, Hofmann^22^, Jentzsch^23^, Musahl^31^). Milrinone showed vasodilatory effects, particularly at higher doses and in combination with nimodipine, but clinical outcomes were inconsistently reported (Duman^19^). Papaverine often produced stronger angiographic dilation than nimodipine without clear clinical advantage (Kerz^25^); bedside continuation of spasmolysis in the ICU was linked to improved outcomes in one historical comparison (Bele^40^).

Angioplasty was widely used as rescue or adjunct therapy. One cohort found no evidence of delayed arterial re-narrowing after angioplasty (Neumann^32^). Device-based strategies, including stent retrievers with nicardipine and permanent stents, were described as technically feasible in refractory cases, but evidence remains limited and highly selected (Kwon^29^, Khanafer^26^).

Standardized perfusion imaging to guide rescue was associated with fewer infarcts and better function in one cohort (Mielke^30^). Others noted discordance between immediate angiographic response and downstream infarction or clinical outcome, and in some cases greater vasodilation on delayed angiography than immediately post-procedure (Schacht^35^, Ott^33^, Zaeske^38^).

Most series reported acceptable safety profiles, however, the reporting and definition of complications were heterogenous. High complication rates and a 6% mortality were noted in one cohort, advising caution in interpretation without controls (Kapapa^24^). Continuous IAN was generally safe yet carried extracranial risks, such as cardiac problems and infections (Kieninger^24^). Physiologic improvements such as normalization of ICP and brain tissue oxygenation were observed during intra-arterial papaverine in patients failing medical therapy (Stiefel^36^).

### Case series

Small single-center series consistently report angiographic improvement after endovascular rescue. For IAN, a 42-patient series with 101 infusions showed 82.2% angiographic response, 68.3% immediate clinical improvement, 21.4% vasospasm-related infarction, and no drug-related complications (Cho^42^). Other reports describe reversal of angiographic spasm with clinical benefit varying by timing, vasospasm burden, and number of procedures (Bashir^39^). Continuous IAN was associated with better outcomes and fewer strokes versus a historical control (Bele^40^). Additional series judged IAN effective and safe in selected patients while calling for prospective confirmation (Biondi^41^).

In a 69-patient comparison, intra-arterial papaverine, balloon angioplasty, or both improved angiography without differences in short-term neurological improvement; effects did not correlate consistently with TCD and showed no clear time-dependence (Coenen^43^).

### Synthesis of results - Meta-Analysis

We planned to pool three outcomes: resolution of angiographic vasospasm, DCI-related infarction, and dichotomized functional outcome. Reporting of the first two was too sparse or inconsistently defined to permit pooling, and too few studies included a control group for comparative meta-analysis. We therefore performed single-arm meta-analyses of proportions for dichotomized functional outcome and present pooled estimates overall and stratified by follow-up time (≤90 vs >90 days), intervention type (intra-arterial spasmolysis, balloon angioplasty, combined), and clinical severity (≥40% vs <40% poor grade).

### Stratified by follow up time

We synthesized single-arm proportions of favorable functional outcome on the logit scale using inverse-variance weighting and fitted both common-effect and random-effects models, reporting back-transformed proportions. These results are presented as a forest plot (**Fig. 2**). Twenty-seven studies (N = 1,175) were included. Overall, the pooled proportion was 0.55 (95% CI 0.49–0.61) under the random-effects model and 0.52 (0.49–0.55) under the common-effect model, with substantial heterogeneity (I² = 71%, τ² = 0.290, p < 0.01). By follow-up, ≤90 days (9 studies; N = 321) yielded 0.58 (0.45–0.70) with high heterogeneity (I² = 80%, τ² = 0.395), and >90 days (18 studies; N = 854) yielded 0.54 (0.47–0.61) with moderate heterogeneity (I² = 65%, τ² = 0.239). Tests for subgroup differences were not statistically significant (random-effects χ² = 0.27, p = 0.60; common-effect χ² = 3.18, p = 0.07). Most precision came from the >90-day subgroup, which contributed about 70% of the total weight.

**Figure 2.**
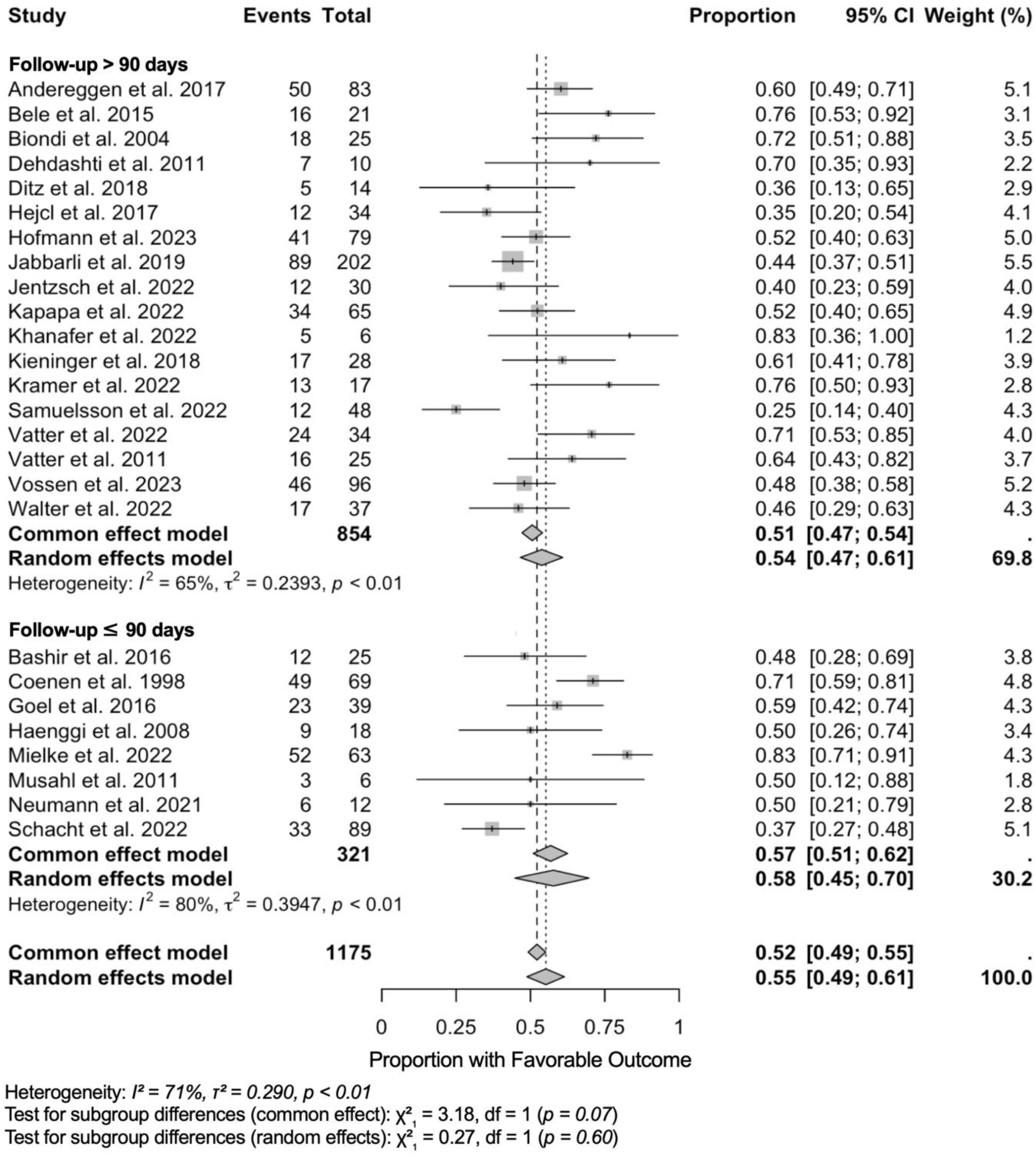
Forest plot of favorable functional outcomes following endovascular treatment for delayed cerebral ischemia, stratified by follow-up duration. Forest plot showing proportions of patients achieving favorable functional outcomes after endovascular treatment for delayed cerebral ischemia (DCI), stratified by follow-up duration. Each study is represented by a square with horizontal lines indicating 95% confidence intervals (CI). Square size is proportional to study weight in the meta-analysis. Studies are grouped by follow-up duration: long-term follow-up (>90 days) and short-term follow-up (≤90 days). Pooled estimates are shown as diamonds for each subgroup and overall. The common effect model assumes homogeneous treatment effects across studies, while the random effects model accounts for between-study heterogeneity. Study weights are calculated using the inverse variance method, where larger studies and those with more precise estimates receive greater weight. Heterogeneity statistics: I² represents the percentage of variability due to heterogeneity rather than sampling error; τ² (tau-squared) quantifies between-study variance; Q-test p-values assess statistical significance of heterogeneity. Subgroup differences were tested using chi-square tests for both common and random effects models. The vertical dashed line at 0.5 represents the null hypothesis of no treatment benefit (50% favorable outcome rate). Results favoring treatment efficacy appear to the right of this line. A total of 26 studies with 1,175 patients and 621 events were included in this analysis. CI, confidence interval; df, degrees of freedom; I^2^, I square statistic providing the percentage of variation across studies that is due to heterogeneity; τ² (tau-squared statistic representing between-study variance in the random effects meta-analysis; χ², (chi-squared) statistic refers to the test statistics for subgroup differences.

### Stratified by endovascular method

We synthesized single-arm proportions of favorable functional outcome on the logit scale using inverse-variance weighting and fit both common-effect and random-effects models, reporting back-transformed proportions. These results are depicted as a forest plot in **Fig. 3**. Across 31 studies (N = 1,377), the pooled proportion was 0.56 (95% CI 0.50–0.61) under the random-effects model and 0.53 (0.50–0.55) under the common-effect model, with substantial heterogeneity (I² = 71%, τ² = 0.283, p < 0.01). By intervention: combined therapy (intra-arterial spasmolysis + balloon angioplasty; N = 692) yielded 0.57 (0.48–0.67) with I² = 78% (τ² = 0.396); intra-arterial spasmolysis alone (N = 444) yielded 0.50 (0.43–0.58) with I² = 57% (τ² = 0.131); balloon angioplasty alone (N = 241) yielded 0.60 (0.47– 0.72) with I² = 74% (τ² = 0.277). Tests for subgroup differences were not statistically significant.

**Figure 3.**
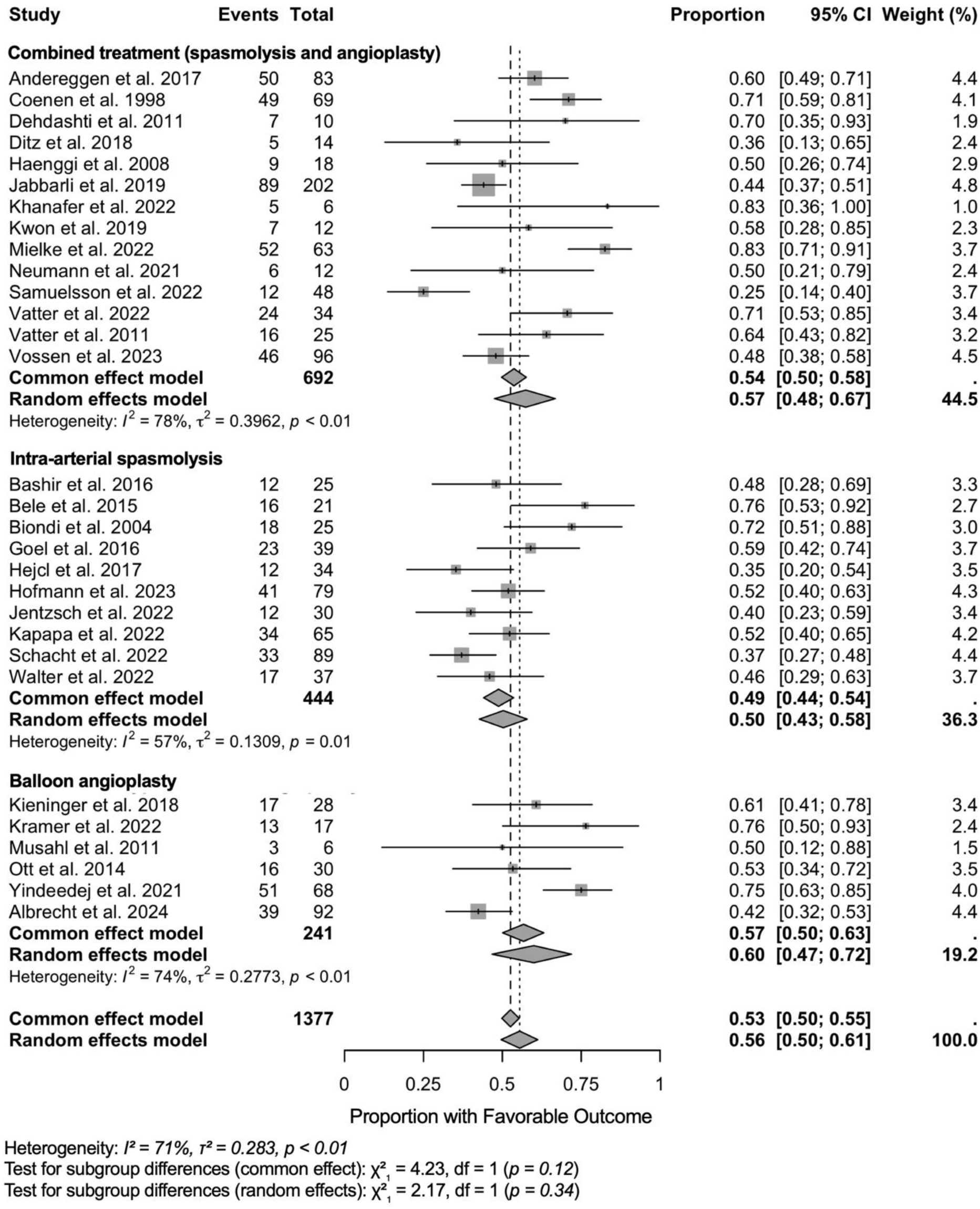
Forest plot of favorable functional outcomes following endovascular treatment for delayed cerebral ischemia, stratified by intervention type. Forest plot showing proportions of patients achieving favorable functional outcomes after endovascular treatment for delayed cerebral ischemia (DCI), stratified by intervention type. Each study is represented by a square with horizontal lines indicating 95% confidence intervals (CI). Square size is proportional to study weight in the meta-analysis. Studies are grouped by intervention type: combined treatment (spasmolysis and angioplasty), intra-arterial spasmolysis alone, and balloon angioplasty alone. Pooled estimates are shown as diamonds for each subgroup and overall. The common effect model assumes homogeneous treatment effects across studies, while the random effects model accounts for between-study heterogeneity. Study weights are calculated using the inverse variance method, where larger studies and those with more precise estimates receive greater weight. Heterogeneity statistics: I² represents the percentage of variability due to heterogeneity rather than sampling error; τ² (tau-squared) quantifies the between-study variance component in the random effects model; Q-test p-values assess statistical significance of heterogeneity. Subgroup differences were tested using chi-square tests: no statistically significant differences were found between intervention types (common effect: χ² = 4.23, p = 0.12; random effects: χ² = 2.17, p = 0.34). The vertical dashed line at 0.5 represents the null hypothesis of no treatment benefit (50% favorable outcome rate). Results favoring treatment efficacy appear to the right of this line. A total of 30 studies with 1,377 patients and 734 events were included in this analysis. CI, confidence interval; df, degrees of freedom; I^2^, I square statistic providing the percentage of variation across studies that is due to heterogeneity; τ² (tau-squared statistic representing between-study variance in the random effects meta-analysis; χ², (chi-squared) statistic refers to the test statistics for subgroup differences.

### Stratified by clinical severity

We synthesized single-arm proportions of favorable functional outcome on the logit scale using inverse-variance weighting and fit both common-effect and random-effects models, reporting back-transformed estimates. These results are graphically depicted as a forest plot in **Fig. 4**. Twenty-six studies (N = 1,140) contributed. Overall, the pooled proportion was 0.54 (95% CI 0.47–0.60) under the random-effects model and 0.50 (0.47–0.53) under the common-effect model, with substantial heterogeneity (I² = 72%, τ² = 0.322, p < 0.01). In the high-severity subgroup (≥40% poor grade; N = 518), the pooled estimate was 0.54 (0.44–0.64) with I² = 79% (τ² = 0.411), contributing ∼51.5% of the total weight. In the low-severity subgroup (<40% poor grade; N = 622), the pooled estimate was 0.52 (0.44–0.61) with I² = 59% (τ² = 0.219). Tests for subgroup differences were not statistically significant (random-effects χ² = 0.11, p = 0.75; common-effect χ² = 3.49, p = 0.06).

**Figure 4.**
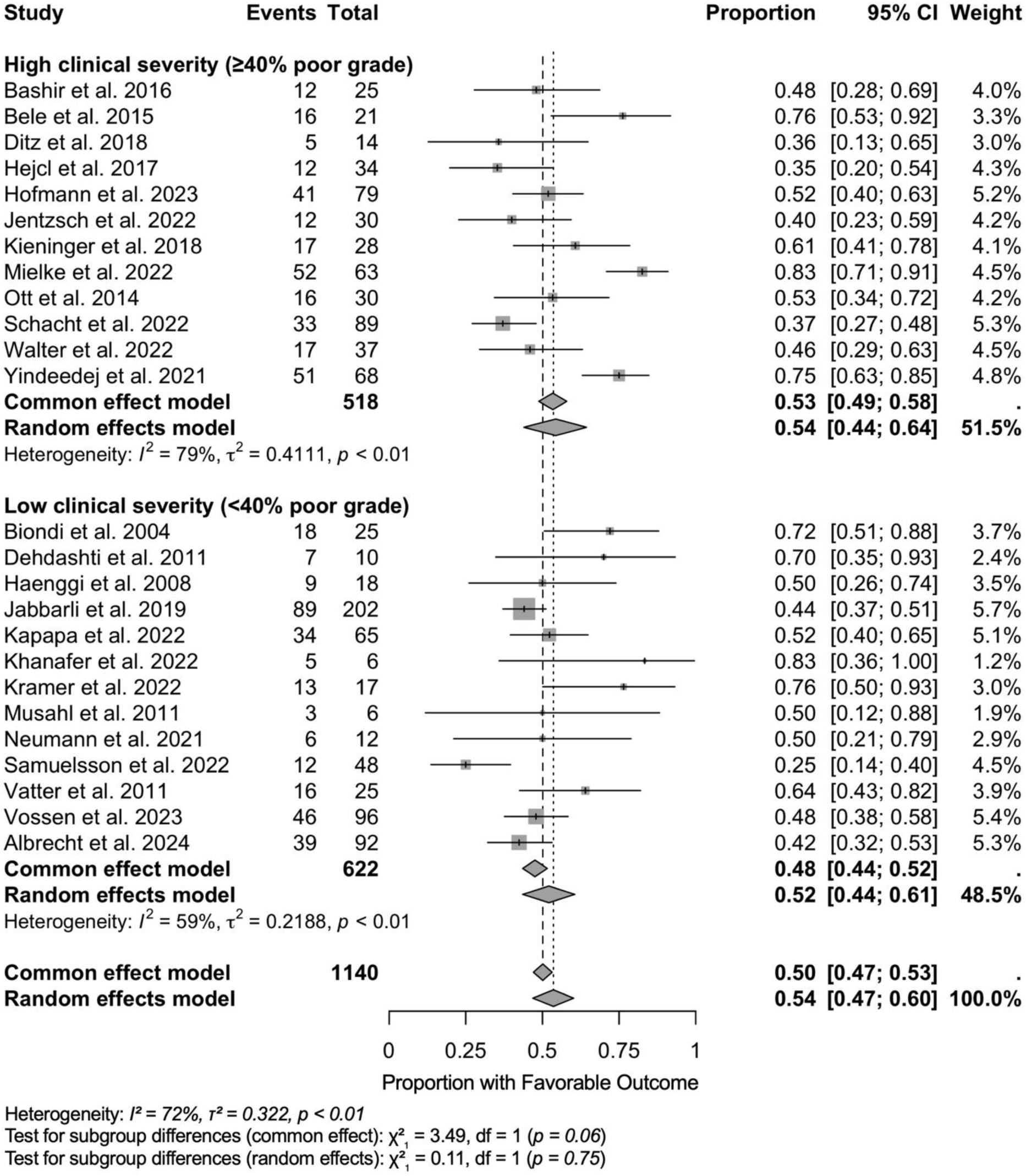
Forest plot of favorable functional outcomes following endovascular treatment for delayed cerebral ischemia, stratified by clinical severity. Forest plot showing proportions of patients achieving favorable functional outcomes after endovascular treatment for delayed cerebral ischemia (DCI), stratified by clinical severity at presentation. For this purpose, poor-grade aSAH is defined as either Hunt & Hess grade 4-5 or WFNS grade 4-5. Each study is represented by a square with horizontal lines indicating 95% confidence intervals (CI). Square size is proportional to study weight in the meta-analysis. Studies are grouped by clinical severity: high clinical severity (≥40% poor grade patients) and low clinical severity (<40% poor grade patients). Pooled estimates are shown as diamonds for each subgroup and overall. The common effect model assumes homogeneous treatment effects across studies, while the random effects model accounts for between-study heterogeneity. Study weights are calculated using the inverse variance method, where larger studies and those with more precise estimates receive greater weight. Heterogeneity statistics: I² represents the percentage of variability due to heterogeneity rather than sampling error; τ² (tau-squared) quantifies the between-study variance component in the random effects model; Q-test p-values assess statistical significance of heterogeneity. Subgroup differences were tested using chi-square tests: no statistically significant differences were found between clinical severity groups (common effect: χ² = 3.49, p = 0.06; random effects: χ² = 0.11, p = 0.75). The vertical dashed line at 0.5 represents the null hypothesis of no treatment benefit (50% favorable outcome rate). Results favoring treatment efficacy appear to the right of this line. A total of 25 studies with 1,140 patients were included in this analysis. Both high and low clinical severity groups demonstrated similar treatment effectiveness, suggesting that endovascular intervention benefits patients regardless of initial clinical severity. CI, confidence interval; df, degrees of freedom; I^2^, I square statistic providing the percentage of variation across studies that is due to heterogeneity; τ² (tau-squared statistic representing between-study variance in the random effects meta-analysis; χ², (chi-squared) statistic refers to the test statistics for subgroup differences.

**Table 1.**
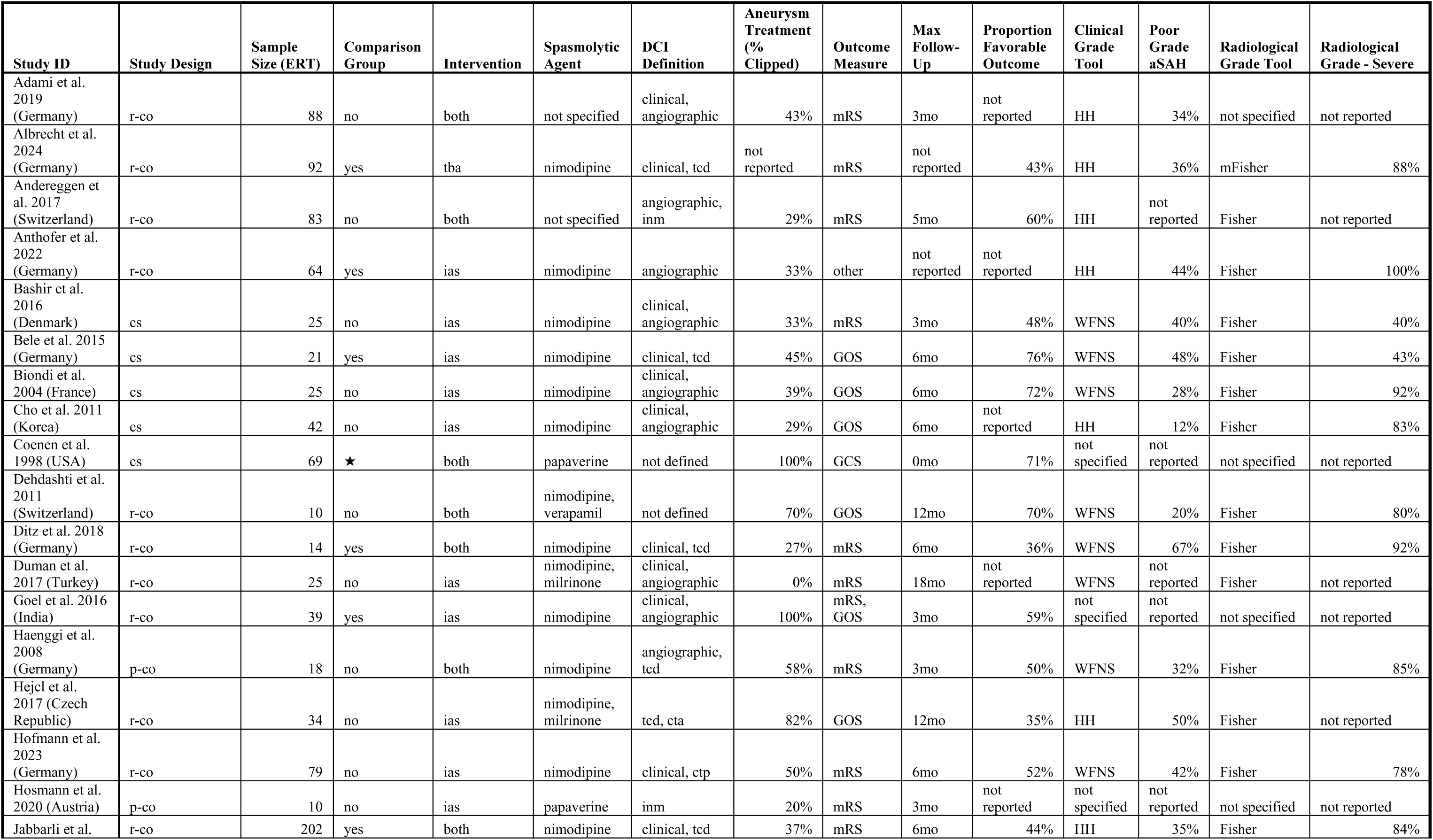
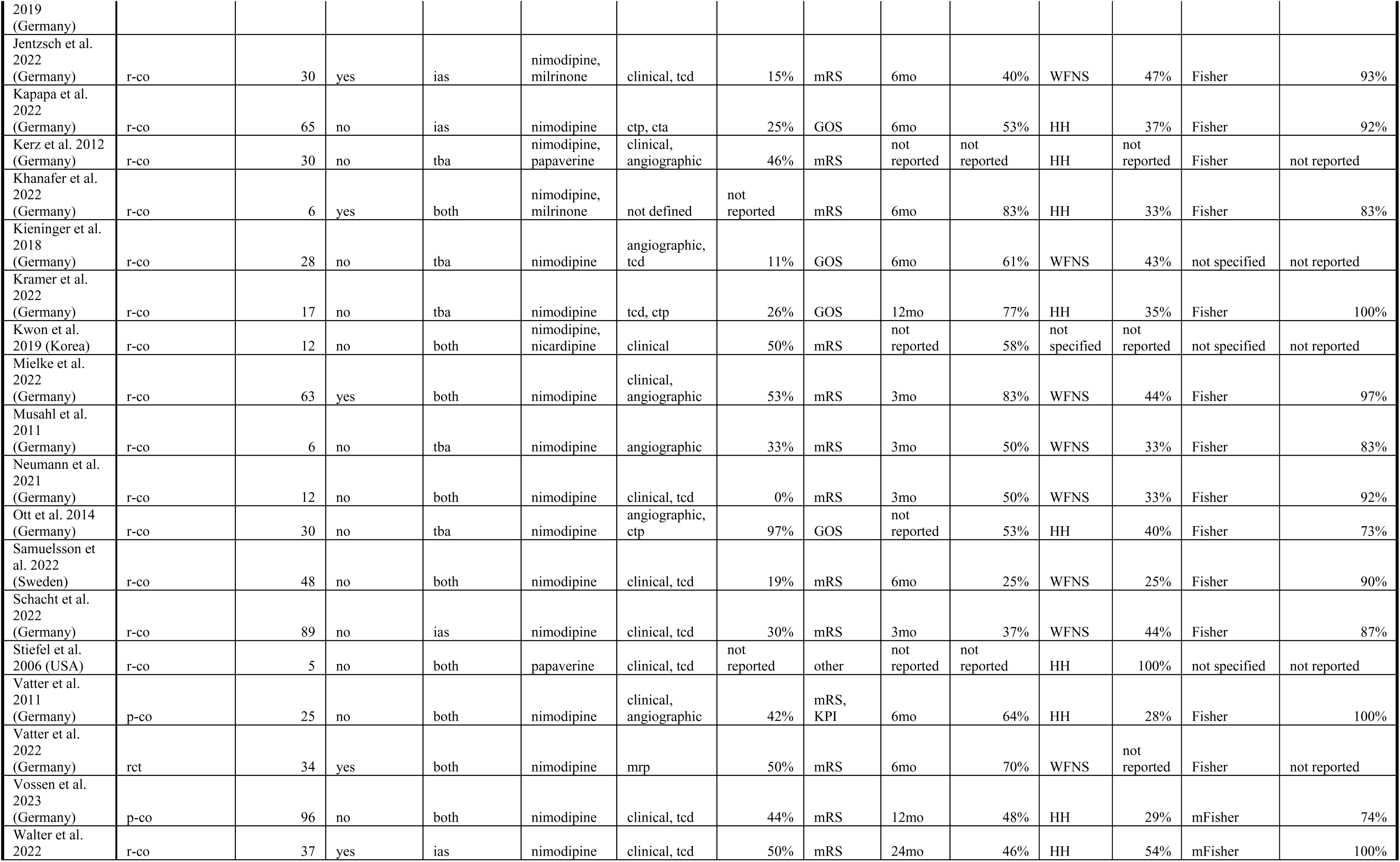
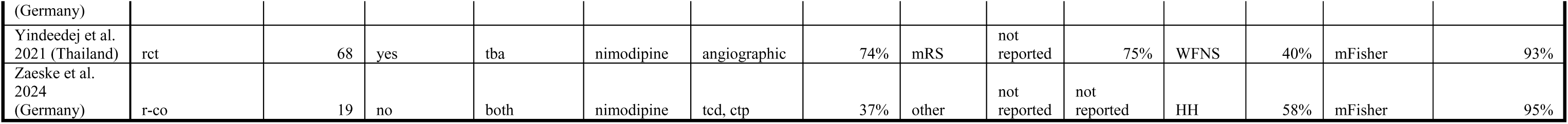
Characteristics of included studies on endovascular treatment for delayed cerebral ischemia. Summary of study characteristics, patient populations, interventions, and outcomes for the 38 studies included in the meta-analysis. Sample sizes reflect only patients who received endovascular treatment for delayed cerebral ischemia (DCI). Control group patients were excluded from analysis due to heterogeneous definitions across studies. Favorable functional outcomes were defined as modified Rankin Scale (mRS) scores of 0-3 or Glasgow Outcome Scale (GOS) scores of 4-5. Poor clinical grade was defined as the proportion of patients presenting with severe neurological status at admission as either Hunt & Hess grade 4-5 or WFNS grade 4-5. Severe radiological grade represents the proportion of patients with Fisher grade 3-4 or modified Fisher grade 3-4 on initial imaging. While pooling these grading scales is not methodologically ideal as they represent slightly different clinical and radiological states, this approach was deemed acceptable as a summary measure while acknowledging its inherent limitations. The percentage of aneurysms treated by surgical clipping is reported as aneurysm securing method, as endovascular aneurysm treatment encompasses diverse techniques (coiling, flow diversion, intra-saccular devices, etc.) that are more difficult to summarize as a single measure. cs, case series; cty, computed tomography angiography; ctp, computed tomography perfusion scanning; Fisher, Fisher grading scale; GOS, Glasgow outcome scale; HH, Hunt & Hess grading; ias, intra-arterial spasmolysis; inm, invasive neuromonitoring; KPI, Karnofsky performance index; mo, months; mrp; magnetic resonance perfusion scanning; mRS, modified Rankin scale; mFisher, modified Fisher grading scale; p-co, prospective cohort study; r-co, retrospective cohort study; rct, randomized controlled trial; aSAH, subarachnoid hemorrhage; tba, transluminal balloon angioplasty; tcd, transcranial Doppler sonsography; WFNS, world federation of neurosurgical societies grading.

| Study ID | Study Design | Sample Size (ERT) | Comparison Group | Intervention | Spasmolytic Agent | DCI Definition | Aneurysm Treatment (% Clipped) | Outcome Measure | Max Follow-Up | Proportion Favorable Outcome | Clinical Grade Tool | Poor Grade aSAH | Radiological Grade Tool | Radiological Grade - Severe |
| --- | --- | --- | --- | --- | --- | --- | --- | --- | --- | --- | --- | --- | --- | --- |
| Adami et al. 2019 (Germany) | r-co | 88 | no | both | not specified | clinical, angiographic | 43% | mRS | 3mo | not reported | HH | 34% | not specified | not reported |
| Albrecht et al. 2024 (Germany) | r-co | 92 | yes | tba | nimodipine | clinical, tcd | not reported | mRS | not reported | 43% | HH | 36% | mFisher | 88% |
| Andereggen et al. 2017 (Switzerland) | r-co | 83 | no | both | not specified | angiographic, inm | 29% | mRS | 5mo | 60% | HH | not reported | Fisher | not reported |
| Anthofer et al. 2022 (Germany) | r-co | 64 | yes | ias | nimodipine | angiographic | 33% | other | not reported | not reported | HH | 44% | Fisher | 100% |
| Bashir et al. 2016 (Denmark) | cs | 25 | no | ias | nimodipine | clinical, angiographic | 33% | mRS | 3mo | 48% | WFNS | 40% | Fisher | 40% |
| Bele et al. 2015 (Germany) | cs | 21 | yes | ias | nimodipine | clinical, tcd | 45% | GOS | 6mo | 76% | WFNS | 48% | Fisher | 43% |
| Biondi et al. 2004 (France) | cs | 25 | no | ias | nimodipine | clinical, angiographic | 39% | GOS | 6mo | 72% | WFNS | 28% | Fisher | 92% |
| Cho et al. 2011 (Korea) | cs | 42 | no | ias | nimodipine | clinical, angiographic | 29% | GOS | 6mo | not reported | HH | 12% | Fisher | 83% |
| Coenen et al. 1998 (USA) | cs | 69 | ★ | both | papaverine | not defined | 100% | GCS | 0mo | 71% | not specified | not reported | not specified | not reported |
| Dehdashti et al. 2011 (Switzerland) | r-co | 10 | no | both | nimodipine, verapamil | not defined | 70% | GOS | 12mo | 70% | WFNS | 20% | Fisher | 80% |
| Ditz et al. 2018 (Germany) | r-co | 14 | yes | both | nimodipine | clinical, tcd | 27% | mRS | 6mo | 36% | WFNS | 67% | Fisher | 92% |
| Duman et al. 2017 (Turkey) | r-co | 25 | no | ias | nimodipine, milrinone | clinical, angiographic | 0% | mRS | 18mo | not reported | WFNS | not reported | Fisher | not reported |
| Goel et al. 2016 (India) | r-co | 39 | yes | ias | nimodipine | clinical, angiographic | 100% | mRS, GOS | 3mo | 59% | not specified | not reported | not specified | not reported |
| Haenggi et al. 2008 (Germany) | p-co | 18 | no | both | nimodipine | angiographic, tcd | 58% | mRS | 3mo | 50% | WFNS | 32% | Fisher | 85% |
| Hejcl et al. 2017 (Czech Republic) | r-co | 34 | no | ias | nimodipine, milrinone | tcd, cta | 82% | GOS | 12mo | 35% | HH | 50% | Fisher | not reported |
| Hofmann et al. 2023 (Germany) | r-co | 79 | no | ias | nimodipine | clinical, ctp | 50% | mRS | 6mo | 52% | WFNS | 42% | Fisher | 78% |
| Hosmann et al. 2020 (Austria) | p-co | 10 | no | ias | papaverine | inm | 20% | mRS | 3mo | not reported | not specified | not reported | not specified | not reported |
| Jabbarli et al. | r-co | 202 | yes | both | nimodipine | clinical, tcd | 37% | mRS | 6mo | 44% | HH | 35% | Fisher | 84% |
| 2019<br>(Germany) |  |  |  |  |  |  |  |  |  |  |  |  |  |  |
| Jentzsch et al.<br>2022<br>(Germany) | r-co | 30 | yes | ias | nimodipine,<br>milrinone | clinical, tcd | 15% | mRS | 6mo | 40% | WFNS | 47% | Fisher | 93% |
| Kapapa et al.<br>2022<br>(Germany) | r-co | 65 | no | ias | nimodipine | ctp, cta | 25% | GOS | 6mo | 53% | HH | 37% | Fisher | 92% |
| Kerz et al. 2012<br>(Germany) | r-co | 30 | no | tba | nimodipine,<br>papaverine | clinical,<br>angiographic | 46% | mRS | not<br>reported | not<br>reported | HH | not<br>reported | Fisher | not reported |
| Khanafer et al.<br>2022<br>(Germany) | r-co | 6 | yes | both | nimodipine,<br>milrinone | not defined | not<br>reported | mRS | 6mo | 83% | HH | 33% | Fisher | 83% |
| Kieninger et al.<br>2018<br>(Germany) | r-co | 28 | no | tba | nimodipine | angiographic,<br>tcd | 11% | GOS | 6mo | 61% | WFNS | 43% | not specified | not reported |
| Kramer et al.<br>2022<br>(Germany) | r-co | 17 | no | tba | nimodipine | tcd, ctp | 26% | GOS | 12mo | 77% | HH | 35% | Fisher | 100% |
| Kwon et al.<br>2019 (Korea) | r-co | 12 | no | both | nimodipine,<br>nicardipine | clinical | 50% | mRS | not<br>reported | 58% | not<br>specified | not<br>reported | not specified | not reported |
| Mielke et al.<br>2022<br>(Germany) | r-co | 63 | yes | both | nimodipine | clinical,<br>angiographic | 53% | mRS | 3mo | 83% | WFNS | 44% | Fisher | 97% |
| Musahl et al.<br>2011<br>(Germany) | r-co | 6 | no | tba | nimodipine | angiographic | 33% | mRS | 3mo | 50% | WFNS | 33% | Fisher | 83% |
| Neumann et al.<br>2021<br>(Germany) | r-co | 12 | no | both | nimodipine | clinical, tcd | 0% | mRS | 3mo | 50% | WFNS | 33% | Fisher | 92% |
| Ott et al. 2014<br>(Germany) | r-co | 30 | no | tba | nimodipine | angiographic,<br>ctp | 97% | GOS | not<br>reported | 53% | HH | 40% | Fisher | 73% |
| Samuelsson et<br>al. 2022<br>(Sweden) | r-co | 48 | no | both | nimodipine | clinical, tcd | 19% | mRS | 6mo | 25% | WFNS | 25% | Fisher | 90% |
| Schacht et al.<br>2022<br>(Germany) | r-co | 89 | no | ias | nimodipine | clinical, tcd | 30% | mRS | 3mo | 37% | WFNS | 44% | Fisher | 87% |
| Stiefel et al.<br>2006 (USA) | r-co | 5 | no | both | papaverine | clinical, tcd | not<br>reported | other | not<br>reported | not<br>reported | HH | 100% | not specified | not reported |
| Vatter et al.<br>2011<br>(Germany) | p-co | 25 | no | both | nimodipine | clinical,<br>angiographic | 42% | mRS,<br>KPI | 6mo | 64% | HH | 28% | Fisher | 100% |
| Vatter et al.<br>2022<br>(Germany) | rct | 34 | yes | both | nimodipine | mrp | 50% | mRS | 6mo | 70% | WFNS | not<br>reported | Fisher | not reported |
| Vossen et al.<br>2023<br>(Germany) | p-co | 96 | no | both | nimodipine | clinical, tcd | 44% | mRS | 12mo | 48% | HH | 29% | mFisher | 74% |
| Walter et al.<br>2022 | r-co | 37 | yes | ias | nimodipine | clinical, tcd | 50% | mRS | 24mo | 46% | HH | 54% | mFisher | 100% |
| (Germany) |  |  |  |  |  |  |  |  |  |  |  |  |  |  |
| Yindeedej et al. 2021 (Thailand) | rct | 68 | yes | tba | nimodipine | angiographic | 74% | mRS | not reported | 75% | WFNS | 40% | mFisher | 93% |
| Zaeske et al. 2024 (Germany) | r-co | 19 | no | both | nimodipine | tcd, ctp | 37% | other | not reported | not reported | HH | 58% | mFisher | 95% |

## Discussion

### Summary of evidence

Taken together, retrospective data indicate that endovascular rescue therapies are frequently associated with angiographic improvement; however, consistent evidence of clinical benefit remains limited. This likely reflects heterogeneity in patient selection, treatment technique and intensity, as well as variation in outcome assessment and timing. The observational nature of most studies, along with missing data and unmatched cohorts, further constrains causal interpretation. These considerations informed our use of random-effects models, stratification by follow-up and intervention type, and a cautious interpretation of pooled results.

Across 39 studies including over 1,600 patients, designs, case mix, interventions, and outcome timing varied widely. Most reports were retrospective, single-center cohorts, and only two small randomized trials provided conflicting short-term results with limited follow-up. Angiographic improvement after endovascular rescue was common, but translation into infarct prevention and durable functional recovery remained inconsistent.

Although DCI-related infarction was one of the principal outcome of interest, heterogeneous definitions and incomplete reporting precluded meaningful pooling or comparison across studies. Consequently, our quantitative synthesis focused on functional outcomes, which showed favorable recovery in roughly half of treated patients, albeit with substantial heterogeneity. Stratified analyses by follow-up duration, intervention type, and clinical severity did not identify clear subgroup differences. Notably, studies employing continuous intra-arterial nimodipine often reported short-term neurological improvement and, in some cases, lower infarction rates compared with conservatively treated cohorts; however, these observations were derived mainly from non-randomized data and remain susceptible to selection and indication biases. Two randomized controlled trials were included. One reported no benefit, and even a potential signal toward harm, with early endovascular rescue therapy. The other suggested a reduction in stroke burden and improvement in short-term neurological outcomes, albeit with limitations in follow-up and patient selection. Across studies with control groups (n = 13), results were mixed. The majority of studies did not provide a control group with best medical treatment alone. Some studies that did, demonstrated clinical benefit while others showed comparable or inferior outcomes compared to medical therapy.

The risk of complications varied across studies. While IAN was generally associated with a favorable safety profile, balloon angioplasty carried a higher rate of procedural complications, including arterial dissection and thromboembolism. Reported complication rates ranged from 3% to 17%, with higher rates associated with mechanical interventions or repeated procedures.

Our findings are amidst prior reviews with different emphases. Ma et al. compared endovascular therapy with standard care and reported lower in-hospital mortality, but no improvement in discharge or follow-up functional outcomes and longer ICU and hospital stays ^48^. Our single-arm proportions are consistent with uncertain functional benefit at follow-up yet cannot address mortality. Boulouis et al., pooling broader CVS-targeted strategies, suggested a relative benefit of intra-arterial approaches in refractory vasospasm ^49^; In this review differences in scope, comparators, and timeframe likely explain the discrepancy with our non-comparative, function-focused estimates. Viderman et al. summarized adverse effects of continuous intra-arterial nimodipine, highlighting hypotension and hematologic complications; this aligns with safety signals noted in several cohorts in our review, although we did not meta-analyze harms ^50^.

### Limitations

Evidence quality was limited. Most studies were observational, small, and single-center, often without concurrent controls, leaving results prone to selection, confounding, and information biases. Outcome scales (mRS vs GOS) and assessment time points varied; despite stratification, clinical and methodological heterogeneity remained high (I² ∼60–80% within strata). Our primary synthesis used single-arm proportions, which describe outcomes after rescue therapy but cannot establish comparative effectiveness versus medical management or optimal treatment sequencing. Reporting of angiographic endpoints, perfusion metrics, and DCI-related infarction was inconsistent, precluding quantitative pooling for these outcomes. Safety reporting was heterogeneous, so complication estimates were summarized narratively rather than pooled; reported rates ranged widely and were higher with mechanical interventions in some series. Risk-of-bias assessment (mean NOS 5.15/9) indicated generally moderate to high risk, with frequent shortcomings in group comparability and follow-up. Finally, intervention categorization was not uniform across studies, and some cohorts combined pharmacologic and mechanical rescue, limiting inference about the relative contribution of specific techniques.

## Conclusion

Across 39 studies, endovascular rescue for delayed cerebral ischemia after aneurysmal subarachnoid hemorrhage was consistently associated with angiographic improvement, yet its impact on functional recovery remained inconsistent. Safety profiles were generally acceptable, although complication reporting varied and some series suggested higher risks with mechanical interventions.

Current evidence does not establish superiority over medical management. Future research should prioritize adequately powered, multicenter randomized trials; standardized criteria for patient selection, timing, and outcome assessment; and transparent reporting of adverse events. Until such data are available, endovascular treatment should be regarded as a rescue option for carefully selected patients, acknowledging the ongoing uncertainty regarding its comparative clinical effectiveness.

## Statements and Declarations

### Authors’ contributions

The design and conception of this trial was developed by MV. Titles and abstracts for potential papers were screened by MV and CSW. Full text screening was caried out by MV, TR, RR and CSW. The meta-analysis was performed by MV, RH and HS. The study was supervised by CSW. The manuscript was drafted and illustrations were created by MV. The final manuscript was critically revised and approved by all authors.

## Declaration of conflicting interest

There are no conflicts of interest to report.

## Funding statement

This project was made possible by the generous funding of the German Research Foundation (Deutsche Forschungsgemeinschaft, DFG), through which Michael Veldeman received a Walter Benjamin Scholarship (Grant Number: VE 1274/1-2). No funding bodies had any involvement in the preparation of this trial or in the decision to submit the paper for publication.

## Abbreviations

AHRQ: Agency for Healthcare Research and Quality
aSAH: Aneurysmal subarachnoid hemorrhage
cs: case series
CI: Confidence interval
CT: Computed tomography
DCI: Delayed cerebral ischemia
GLMM: Generalized linear mixed effects model
GOS: Glasgow outcome scale
ICU: intensive care unit
mRS: modified Ranking scale
NOS: Newcastle–Ottawa Scale
p-co: Prospective cohort study
r-co: Retrospective cohort study
RCT: randomized controlled trial
RoB: Risk of bias

## Data Availability

This being a Systematic Review and Meta Analysis, all data is available in the manuscript its tables and the source articles on which it is based.

---

**Supplemental figure 1.**
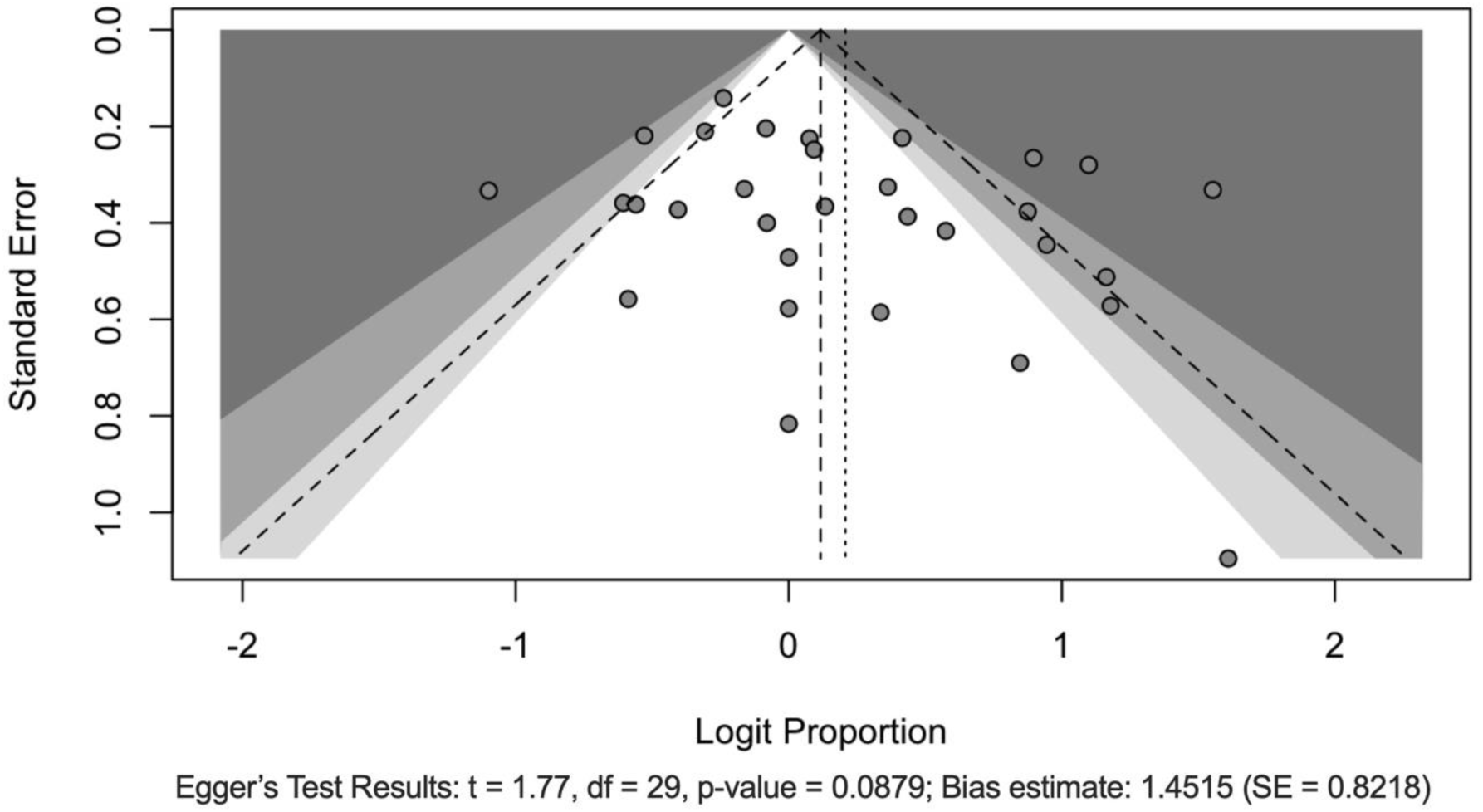
Funnel plot for assessment of publication bias in studies of endovascular treatment for delayed cerebral ischemia. Funnel plot showing the relationship between study precision (standard error) and effect size (logit-transformed proportions of favorable functional outcomes). Each circle represents an individual study. The vertical dashed line indicates the overall pooled effect estimate from the random effects meta-analysis. The diagonal dashed lines represent the 95% confidence limits around the pooled estimate, creating a triangular region where studies would be expected to fall in the absence of publication bias. Egger’s regression test was performed to statistically assess funnel plot asymmetry (t = 1.77, df = 29, p = 0.0879), indicating no statistically significant evidence of publication bias at the conventional α = 0.05 level, though the p-value approaches significance suggesting potential small-study effects.

**Supplementary Table 1.** Risk of bias assessment using the Newcastle-Ottawa Scale for non-randomized studies. Risk of bias assessment for included studies using the Newcastle-Ottawa Scale (NOS), with scores ranging from 0-8 stars across eight domains. Overall quality ratings were assigned according to Agency for Healthcare Research and Quality (AHRQ) standards: High quality (7-8 stars, low risk of bias), Moderate quality (5-6 stars, moderate risk of bias), and Poor quality (0-4 stars, high risk of bias). **Domain definitions adapted for endovascular DCI treatment studies:** **Representativeness:** Studies reporting the total number of aSAH patients during the inclusion period and selection criteria for endovascular treatment, or describing a consecutive patient series. Either criterion sufficed for a star. **Selection Non-exposed:** Clear definition of endovascular treatment indications specifying patient eligibility criteria. Studies with well-described control groups or adequate baseline characteristics of the source cohort received a star, as most studies lacked control groups. **Ascertainment of Exposure:** Well-described endovascular vasospasm treatment protocols. Studies with vague descriptions of treatment type (spasmolysis vs. angioplasty) received no star. **Outcome Not Present at Start:** This domain was considered inherently satisfied for all studies as patients with DCI represent an acute condition requiring intervention. **Comparability:** Attempts to control for confounding variables between treatment groups. Only applicable to studies with some form of control group. **Outcome Assessment:** Objective outcome measures with direct patient relevance. Studies reporting infarction rates and/or clinical outcomes at discharge or follow-up received a star. Angiographic resolution alone or changes in invasive neuromonitoring were insufficient. **Length of Follow-up:** Minimum 3-month follow-up with functional outcome assessment, preferably using modified Rankin Scale. Clinical outcomes at discharge alone were insufficient. **Adequacy of Follow-up:** Standardized follow-up methodology with documented clinical evaluation or structured telephone interviews for ordinal outcome assessment. Reconstruction from medical records alone received no star. Stars were assigned as specified by the tool coded as: ★ = criterion met; ⋆ = criterion not met. AHRQ, Assessing the Risk of Bias of Individual Studies in Systematic Reviews of Health Care Interventions

| Study ID | Representativeness | Selection Non-Exposed | Ascertainment of Exposure | Outcome not Present at Start | Comparability | Outcome Assessment | Length of Follow-up | Adequacy of Follow-up | Overall Score | AHRQ |
| --- | --- | --- | --- | --- | --- | --- | --- | --- | --- | --- |
| Adami et al. 2019 | ☆ | ☆ | ☆ | ★ | ☆ | ★ | ★ | ☆ | 3 | Poor |
| Albrecht et al. 2024 | ★ | ★ | ★ | ★ | ☆ | ★ | ★ | ★ | 7 | High |
| Andereggen et al. 2017 | ★ | ★ | ☆ | ★ | ☆ | ★ | ★ | ★ | 6 | Moderate |
| Anthofer et al. 2022 | ★ | ★ | ★ | ★ | ☆ | ☆ | ☆ | ★ | 5 | Moderate |
| Bashir et al. 2016 | ☆ | ☆ | ★ | ★ | ☆ | ★ | ★ | ★ | 5 | Moderate |
| Bele et al. 2015 | ☆ | ☆ | ★ | ★ | ☆ | ★ | ★ | ☆ | 4 | Poor |
| Biondi et al. 2004 | ★ | ★ | ★ | ★ | ☆ | ★ | ★ | ☆ | 6 | Moderate |
| Cho et al. 2011 | ★ | ★ | ★ | ★ | ☆ | ★ | ★ | ☆ | 6 | Moderate |
| Coenen et al. 1998 | ★ | ☆ | ★ | ★ | ☆ | ★ | ☆ | ☆ | 4 | Poor |
| Dehdashti et al. 2011 | ☆ | ★ | ☆ | ★ | ☆ | ★ | ★ | ☆ | 4 | Poor |
| Ditz et al. 2018 | ★ | ★ | ★ | ★ | ★ | ★ | ★ | ☆ | 7 | High |
| Duman et al. 2017 | ★ | ☆ | ★ | ★ | ☆ | ★ | ★ | ☆ | 5 | Moderate |
| Goel et al. 2016 | ★ | ★ | ★ | ★ | ☆ | ★ | ★ | ☆ | 6 | Moderate |
| Haenggi et al. 2008 | ★ | ★ | ★ | ★ | ☆ | ★ | ★ | ☆ | 6 | Moderate |
| Hejcl et al. 2017 | ★ | ☆ | ★ | ★ | ☆ | ★ | ★ | ☆ | 5 | Moderate |
| Hofmann et al. 2023 | ★ | ★ | ★ | ★ | ☆ | ★ | ★ | ☆ | 6 | Moderate |
| Hosmann et al. 2020 | ★ | ☆ | ★ | ★ | ☆ | ★ | ★ | ☆ | 5 | Moderate |
| Jabbarli et al. 2019 | ★ | ★ | ★ | ★ | ★ | ★ | ★ | ★ | 8 | High |
| Jentzsch et al. 2022 | ☆ | ☆ | ★ | ★ | ☆ | ★ | ★ | ★ | 5 | Moderate |
| Kapapa et al. 2022 | ★ | ★ | ★ | ★ | ☆ | ★ | ★ | ★ | 7 | High |
| Kerz et al. 2012 | ★ | ☆ | ★ | ★ | ☆ | ☆ | ☆ | ☆ | 3 | Poor |
| Khanafer et al. 2022 | ★ | ☆ | ☆ | ★ | ☆ | ★ | ★ | ★ | 5 | Moderate |
| Kieninger et al. 2018 | ★ | ☆ | ★ | ★ | ☆ | ★ | ★ | ★ | 6 | Moderate |
| Kramer et al. 2022 | ★ | ★ | ★ | ★ | ☆ | ★ | ★ | ★ | 7 | High |
| Kwon et al. 2019 | ☆ | ☆ | ★ | ★ | ☆ | ☆ | ☆ | ☆ | 2 | Poor |
| Mielke et al. 2022 | ★ | ★ | ★ | ★ | ★ | ★ | ★ | ★ | 8 | High |
| Musahl et al. 2011 | ☆ | ☆ | ★ | ★ | ☆ | ★ | ★ | ★ | 5 | Moderate |
| Neumann et al. 2021 | ☆ | ☆ | ★ | ★ | ☆ | ★ | ★ | ★ | 5 | Moderate |
| Ott et al. 2014 | ☆ | ☆ | ★ | ★ | ☆ | ☆ | ☆ | ☆ | 2 | Poor |
| Samuelsson et al. | ☆ | ☆ | ★ | ★ | ☆ | ★ | ★ | ☆ | 4 | Poor |
| 2022 |  |  |  |  |  |  |  |  |  |  |
| Schacht et al. 2022 | ☆ | ☆ | ★ | ★ | ★ | ★ | ★ | ☆ | 5 | Moderate |
| Stiefel et al. 2006 | ☆ | ☆ | ☆ | ★ | ☆ | ☆ | ☆ | ☆ | 1 | Poor |
| Vatter et al. 2011 | ☆ | ★ | ★ | ★ | ☆ | ★ | ★ | ☆ | 5 | Moderate |
| Vatter et al. 2022 | ★ | ★ | ★ | ★ | ★ | ★ | ★ | ★ | 8 | High |
| Vossen et al. 2023 | ★ | ★ | ★ | ★ | ☆ | ★ | ★ | ★ | 7 | High |
| Walter et al. 2022 | ★ | ★ | ★ | ★ | ☆ | ★ | ★ | ☆ | 6 | Moderate |
| Weiss et al. 2019 | ☆ | ☆ | ★ | ★ | ☆ | ★ | ★ | ★ | 5 | Moderate |
| Yindeedej et al. 2021 | ☆ | ☆ | ★ | ★ | ★ | ★ | ☆ | ☆ | 4 | Poor |
| Zaeske et al. 2024 | ☆ | ★ | ★ | ★ | ☆ | ☆ | ☆ | ☆ | 3 | Poor |

## Notes

### Competing Interest Statement

The authors have declared no competing interest.

### Author Declarations

Not applicable for a systematic review and meta analysis

## References

1. Claassen J, Park S. Spontaneous subarachnoid haemorrhage. Lancet. 2022;400:846–862. doi: 10.1016/s0140-6736(22)00938-2

2. Allen GS, Ahn HS, Preziosi TJ, Battye R, Boone SC, Boone SC, Chou SN, Kelly DL, Weir BK, Crabbe RA, et al. Cerebral arterial spasm--a controlled trial of nimodipine in patients with subarachnoid hemorrhage. N Engl J Med. 1983;308:619–624. doi: 10.1056/nejm198303173081103

3. Pickard JD, Murray GD, Illingworth R, Shaw MD, Teasdale GM, Foy PM, Humphrey PR, Lang DA, Nelson R, Richards P, et al. Effect of oral nimodipine on cerebral infarction and outcome after subarachnoid haemorrhage: British aneurysm nimodipine trial. Bmj. 1989;298:636–642. doi: 10.1136/bmj.298.6674.636

4. Gathier CS, van den Bergh WM, van der Jagt M, Verweij BH, Dankbaar JW, Müller MC, Oldenbeuving AW, Rinkel GJE, Slooter AJC. Induced Hypertension for Delayed Cerebral Ischemia After Aneurysmal Subarachnoid Hemorrhage: A Randomized Clinical Trial. Stroke. 2018;49:76–83. doi: 10.1161/strokeaha.117.017956

5. Dodd WS, Laurent D, Dumont AS, Hasan DM, Jabbour PM, Starke RM, Hosaka K, Polifka AJ, Hoh BL, Chalouhi N. Pathophysiology of Delayed Cerebral Ischemia After Subarachnoid Hemorrhage: A Review. Journal of the American Heart Association. 2021;10:e021845. doi: doi:10.1161/JAHA.121.021845

6. Schupper AJ, Eagles ME, Neifert SN, Mocco J, Macdonald RL. Lessons from the CONSCIOUS-1 Study. J Clin Med. 2020;9. doi: 10.3390/jcm9092970

7. Guenego A, Fahed R, Rouchaud A, Walker G, Faizy TD, Sporns PB, Aggour M, Jabbour P, Alexandre AM, Mosimann PJ, et al. Diagnosis and endovascular management of vasospasm after aneurysmal subarachnoid hemorrhage - survey of real-life practices. J Neurointerv Surg. 2024;16:677–683. doi: 10.1136/jnis-2023-020544

8. Jabbarli R, Pierscianek D, Rölz R, Darkwah Oppong M, Kaier K, Shah M, Taschner C, Mönninghoff C, Urbach H, Beck J, et al. Endovascular treatment of cerebral vasospasm after subarachnoid hemorrhage: More is more. Neurology. 2019;93:e458–e466. doi: 10.1212/wnl.0000000000007862

9. Vossen LV, Weiss M, Albanna W, Conzen-Dilger C, Schulze-Steinen H, Rossmann T, Schmidt TP, Höllig A, Wiesmann M, Clusmann H, et al. Intra-arterial nimodipine for the treatment of refractory delayed cerebral ischemia after aneurysmal subarachnoid hemorrhage. J Neurointerv Surg. 2024;17:e31–e40. doi: 10.1136/jnis-2023-021151

10. Vatter H, Güresir E, König R, Durner G, Kalff R, Schuss P, Mayer TE, Konczalla J, Hattingen E, Seifert V, et al. Invasive Diagnostic and Therapeutic Management of Cerebral VasoSpasm after Aneurysmal Subarachnoid Hemorrhage (IMCVS)-A Phase 2 Randomized Controlled Trial. J Clin Med. 2022;11. doi: 10.3390/jcm11206197

11. Yindeedej V, Nimmannitya P, Noiphithak R, Punyarat P, Tantongtip D. Clinical Outcome in Cerebral Vasospasm Patients Treated with and without Intra-Arterial Nimodipine Infusion. J Neurol Surg A Cent Eur Neurosurg. 2022;83:420–426. doi: 10.1055/s-0041-1735860

12. Wells G, Shea B, O’Connell J. The Newcastle-Ottawa Scale (NOS) for Assessing The Quality of Nonrandomised Studies in Meta-analyses. Ottawa Health Research Institute Web site. 2014;7.

13. Adami D, Berkefeld J, Platz J, Konczalla J, Pfeilschifter W, Weidauer S, Wagner M. Complication rate of intraarterial treatment of severe cerebral vasospasm after subarachnoid hemorrhage with nimodipine and percutaneous transluminal balloon angioplasty: Worth the risk? J Neuroradiol. 2019;46:15–24. doi: 10.1016/j.neurad.2018.04.001

14. Albrecht C, Liang R, Trost D, Hostettler I, Renz M, Meyer B, Zimmer C, Kirschke J, Maegerlein C, Bodden J, et al. Endovascular therapy for cerebral vasospasm after aneurysmal subarachnoid hemorrhage: Single-center experience in a high-volume neurovascular unit. Brain Spine. 2024;4:104133. doi: 10.1016/j.bas.2024.104133

15. Andereggen L, Beck J, Z’Graggen WJ, Schroth G, Andres RH, Murek M, Haenggi M, Reinert M, Raabe A, Gralla J. Feasibility and Safety of Repeat Instant Endovascular Interventions in Patients with Refractory Cerebral Vasospasms. AJNR Am J Neuroradiol. 2017;38:561–567. doi: 10.3174/ajnr.A5024

16. Anthofer J, Bele S, Wendl C, Kieninger M, Zeman F, Bruendl E, Schmidt NO, Schebesch KM. Continuous intra-arterial nimodipine infusion as rescue treatment of severe refractory cerebral vasospasm after aneurysmal subarachnoid hemorrhage. J Clin Neurosci. 2022;96:163–171. doi: 10.1016/j.jocn.2021.10.028

17. Dehdashti AR, Binaghi S, Uske A, Regli L. Intraarterial nimodipine for the treatment of symptomatic vasospasm after aneurysmal subarachnoid hemorrhage: a preliminary study. Neurol India. 2011;59:810–816. doi: 10.4103/0028-3886.91356

18. Ditz C, Neumann A, Wojak J, Smith E, Gliemroth J, Tronnier V, Küchler J. Repeated Endovascular Treatments in Patients with Recurrent Cerebral Vasospasms After Subarachnoid Hemorrhage: A Worthwhile Strategy? World Neurosurg. 2018;112:e791–e798. doi: 10.1016/j.wneu.2018.01.156

19. Duman E, Karakoç F, Pinar HU, Dogan R, Fýrat A, Yýldýrým E. Higher dose intra-arterial milrinone and intra-arterial combined milrinone-nimodipine infusion as a rescue therapy for refractory cerebral vasospasm. Interv Neuroradiol. 2017;23:636–643. doi: 10.1177/1591019917732288

20. Goel R, Aggarwal A, Salunke P, Kumar A, Chhabra R. Is intra arterial nimodipine really beneficial in vasospasm following aneurysmal subarachnoid haemorrhage? Br J Neurosurg. 2016;30:407–410. doi: 10.3109/02688697.2016.1161172

21. Hejèl A, Cihláø F, Smolka V, Vachata P, Bartoš R, Procházka J, Cihlář J, Sameš M. Chemical angioplasty with spasmolytics for vasospasm after subarachnoid hemorrhage. Acta Neurochir (Wien). 2017;159:713–720. doi: 10.1007/s00701-017-3104-5

22. Hofmann BB, Karadag C, Rubbert C, Schieferdecker S, Neyazi M, Abusabha Y, Fischer I, Boogaarts HD, Muhammad S, Beseoglu K, et al. Novel Insights into Pathophysiology of Delayed Cerebral Ischemia: Effects of Current Rescue Therapy on Microvascular Perfusion Heterogeneity. Biomedicines. 2023;11. doi: 10.3390/biomedicines11102624

23. Jentzsch J, Ziganshyna S, Lindner D, Merkel H, Mucha S, Schob S, Quäschling U, Hoffmann KT, Werdehausen R, Halama D, et al. Nimodipine vs. Milrinone - Equal or Complementary Use? A Retrospective Analysis. Front Neurol. 2022;13:939015. doi: 10.3389/fneur.2022.939015

24. Kapapa T, König R, Mayer B, Braun M, Schmitz B, Müller S, Schick J, Wirtz CR, Pala A. Adverse Events and Complications in Continuous Intra-arterial Nimodipine Infusion Therapy After Aneurysmal Subarachnoid Hemorrhage. Front Neurol. 2021;12:812898. doi: 10.3389/fneur.2021.812898

25. Kerz T, Boor S, Beyer C, Welschehold S, Schuessler A, Oertel J. Effect of intraarterial papaverine or nimodipine on vessel diameter in patients with cerebral vasospasm after subarachnoid hemorrhage. Br J Neurosurg. 2012;26:517–524. doi: 10.3109/02688697.2011.650737

26. Khanafer A, Cimpoca A, Bhogal P, Bäzner H, Ganslandt O, Henkes H. Intracranial stenting as a bail-out option for posthemorrhagic cerebral vasospasm: a single-center experience with long-term follow-up. BMC Neurol. 2022;22:351. doi: 10.1186/s12883-022-02862-4

27. Kieninger M, Flessa J, Lindenberg N, Bele S, Redel A, Schneiker A, Schuierer G, Wendl C, Graf B, Silbereisen V. Side Effects of Long-Term Continuous Intra-arterial Nimodipine Infusion in Patients with Severe Refractory Cerebral Vasospasm after Subarachnoid Hemorrhage. Neurocrit Care. 2018;28:65–76. doi: 10.1007/s12028-017-0428-1

28. Kramer A, Selbach M, Kerz T, Neulen A, Brockmann MA, Ringel F, Brockmann C. Continuous Intraarterial Nimodipine Infusion for the Treatment of Delayed Cerebral Ischemia After Aneurysmal Subarachnoid Hemorrhage: A Retrospective, Single-Center Cohort Trial. Front Neurol. 2022;13:829938. doi: 10.3389/fneur.2022.829938

29. Kwon HJ, Lim JW, Koh HS, Park B, Choi SW, Kim SH, Youm JY, Song SH. Stent-Retriever Angioplasty for Recurrent Post-Subarachnoid Hemorrhagic Vasospasm - A Single Center Experience with Long-Term Follow-Up. Clin Neuroradiol. 2019;29:751–761. doi: 10.1007/s00062-018-0711-3

30. Mielke D, Döring K, Behme D, Psychogios MN, Rohde V, Malinova V. The Impact of Endovascular Rescue Therapy on the Clinical and Radiological Outcome After Aneurysmal Subarachnoid Hemorrhage: A Safe and Effective Treatment Option for Hemodynamically Relevant Vasospasm? Front Neurol. 2022;13:838456. doi: 10.3389/fneur.2022.838456

31. Musahl C, Henkes H, Vajda Z, Coburger J, Hopf N. Continuous local intra-arterial nimodipine administration in severe symptomatic vasospasm after subarachnoid hemorrhage. Neurosurgery. 2011;68:1541–1547; discussion 1547. doi: 10.1227/NEU.0b013e31820edd46

32. Neumann A, Küchler J, Ditz C, Krajewski K, Leppert J, Schramm P, Schacht H. Non-compliant and compliant balloons for endovascular rescue therapy of cerebral vasospasm after spontaneous subarachnoid haemorrhage: experiences of a single-centre institution with radiological follow-up of the treated vessel segments. Stroke Vasc Neurol. 2021;6:16–24. doi: 10.1136/svn-2020-000410

33. Ott S, Jedlicka S, Wolf S, Peter M, Pudenz C, Merker P, Schürer L, Lumenta CB. Continuous selective intra-arterial application of nimodipine in refractory cerebral vasospasm due to aneurysmal subarachnoid hemorrhage. Biomed Res Int. 2014;2014:970741. doi: 10.1155/2014/970741

34. Samuelsson J, Sunila M, Rentzos A, Nilsson D. Intra-arterial nimodipine for severe cerebral vasospasm after aneurysmal subarachnoid haemorrhage - neurological and radiological outcome. Neuroradiol J. 2022;35:213–219. doi: 10.1177/19714009211036695

35. Schacht H, Küchler J, Neumann A, Schramm P, Tronnier VM, Ditz C. Analysis of Angiographic Treatment Response to Intra-Arterial Nimodipine Bolus Injection in Patients with Medically Refractory Cerebral Vasospasm After Spontaneous Subarachnoid Hemorrhage. World Neurosurg. 2022;162:e457–e467. doi: 10.1016/j.wneu.2022.03.033

36. Stiefel MF, Spiotta AM, Udoetuk JD, Maloney-Wilensky E, Weigele JB, Hurst RW, LeRoux PD. Intra-arterial papaverine used to treat cerebral vasospasm reduces brain oxygen. Neurocrit Care. 2006;4:113–118. doi: 10.1385/ncc:4:2:113

37. Walter J, Grutza M, Möhlenbruch M, Vollherbst D, Vogt L, Unterberg A, Zweckberger K. The Local Intraarterial Administration of Nimodipine Might Positively Affect Clinical Outcome in Patients with Aneurysmal Subarachnoid Hemorrhage and Delayed Cerebral Ischemia. J Clin Med. 2022;11. doi: 10.3390/jcm11072036

38. Zaeske C, Zopfs D, Laukamp K, Lennartz S, Kottlors J, Goertz L, Stetefeld H, Hof M, Abdullayev N, Kabbasch C, et al. Immediate angiographic control after intra-arterial nimodipine administration underestimates the vasodilatory effect. Sci Rep. 2024;14:6154. doi: 10.1038/s41598-024-56807-7

39. Bashir A, Andresen M, Bartek J, Jr., Cortsen M, Eskesen V, Wagner A. Intra-arterial nimodipine for cerebral vasospasm after subarachnoid haemorrhage: Influence on clinical course and predictors of clinical outcome. Neuroradiol J. 2016;29:72–81. doi: 10.1177/1971400915626429

40. Bele S, Proescholdt MA, Hochreiter A, Schuierer G, Scheitzach J, Wendl C, Kieninger M, Schneiker A, Bründl E, Schödel P, et al. Continuous intra-arterial nimodipine infusion in patients with severe refractory cerebral vasospasm after aneurysmal subarachnoid hemorrhage: a feasibility study and outcome results. Acta Neurochir (Wien). 2015;157:2041–2050. doi: 10.1007/s00701-015-2597-z

41. Biondi A, Ricciardi GK, Puybasset L, Abdennour L, Longo M, Chiras J, Van Effenterre R. Intra-arterial nimodipine for the treatment of symptomatic cerebral vasospasm after aneurysmal subarachnoid hemorrhage: preliminary results. AJNR Am J Neuroradiol. 2004;25:1067–1076.

42. Cho WS, Kang HS, Kim JE, Kwon OK, Oh CW, Son YJ, Know BJ, Jung C, Hang MH. Intra-arterial nimodipine infusion for cerebral vasospasm in patients with aneurysmal subarachnoid hemorrhage. Interv Neuroradiol. 2011;17:169–178. doi: 10.1177/159101991101700205

43. Coenen VA, Hansen CA, Kassell NF, Polin RS. Endovascular treatment for symptomatic cerebral vasospasm after subarachnoid hemorrhage: transluminal balloon angioplasty compared with intraarterial papaverine. Neurosurg Focus. 1998;5:e6. doi: 10.3171/foc.1998.5.4.9

44. Hänggi D, Turowski B, Beseoglu K, Yong M, Steiger HJ. Intra-arterial nimodipine for severe cerebral vasospasm after aneurysmal subarachnoid hemorrhage: influence on clinical course and cerebral perfusion. AJNR Am J Neuroradiol. 2008;29:1053–1060. doi: 10.3174/ajnr.A1005

45. Hosmann A, Wang WT, Dodier P, Bavinzski G, Engel A, Herta J, Plöchl W, Reinprecht A, Gruber A. The Impact of Intra-Arterial Papaverine-Hydrochloride on Cerebral Metabolism and Oxygenation for Treatment of Delayed-Onset Post-Subarachnoid Hemorrhage Vasospasm. Neurosurgery. 2020;87:712–719. doi: 10.1093/neuros/nyz500

46. Vatter H, Güresir E, Berkefeld J, Beck J, Raabe A, du Mesnil de Rochemont R, Seifert V, Weidauer S. Perfusion-diffusion mismatch in MRI to indicate endovascular treatment of cerebral vasospasm after subarachnoid haemorrhage. J Neurol Neurosurg Psychiatry. 2011;82:876–883. doi: 10.1136/jnnp.2010.219592

47. Weiss M, Conzen C, Mueller M, Wiesmann M, Clusmann H, Albanna W, Schubert GA. Endovascular Rescue Treatment for Delayed Cerebral Ischemia After Subarachnoid Hemorrhage Is Safe and Effective. Front Neurol. 2019;10:136. doi: 10.3389/fneur.2019.00136

48. Ma YH, Shang R, Li SH, Wang T, Lin S, Zhang CW. Efficacy of endovascular therapy for cerebral vasospasm following aneurysmal subarachnoid hemorrhage: a systematic review and meta-analysis. Front Neurol. 2024;15:1360511. doi: 10.3389/fneur.2024.1360511

49. Boulouis G, Labeyrie MA, Raymond J, Rodriguez-Régent C, Lukaszewicz AC, Bresson D, Ben Hassen W, Trystram D, Meder JF, Oppenheim C, et al. Treatment of cerebral vasospasm following aneurysmal subarachnoid haemorrhage: a systematic review and meta-analysis. Eur Radiol. 2017;27:3333–3342. doi: 10.1007/s00330-016-4702-y

50. Viderman D, Sarria-Santamera A, Bilotta F. Side effects of continuous intra-arterial infusion of nimodipine for management of resistant cerebral vasospasm in subarachnoid hemorrhage patients: A systematic review. Neurochirurgie. 2021;67:461–469. doi: 10.1016/j.neuchi.2021.02.005

